# Modeling physician variability to prioritize relevant medical record information

**DOI:** 10.1101/2020.09.18.20197434

**Authors:** Mohammadamin Tajgardoon, Gregory F Cooper, Andrew J King, Gilles Clermont, Harry Hochheiser, Milos Hauskrecht, Dean F Sittig, Shyam Visweswaran

## Abstract

**Objective:** Patient information can be retrieved more efficiently in electronic medical record (EMR) systems by using machine learning models that predict which information a physician will seek in a clinical context. However, information-seeking behavior varies across EMR users. To explicitly account for this variability, we derived hierarchical models and compared their performance to non-hierarchical models in identifying relevant patient information in intensive care unit (ICU) cases.

**Materials and Methods:** Critical care physicians reviewed ICU patient cases and selected data items relevant for presenting at morning rounds. Using patient EMR data as predictors, we derived hierarchical logistic regression (HLR) and standard logistic regression (LR) models to predict their relevance.

**Results:** In 73 pairs of HLR and LR models, the HLR models achieved an area under the ROC curve of 0.81, 95% CI [0.80, 0.82], which was statistically significantly higher than that of LR models (0.75, 95% CI [0.74-0.76]). Further, the HLR models achieved statistically significantly lower expected calibration error (0.07, 95% CI [0.06-0.08]) than LR models (0.16, 95% CI [0.14-0.17]).

**Discussion:** The physician reviewers demonstrated variability in selecting relevant data. Our results show that HLR models perform significantly better than LR models with respect to both discrimination and calibration. This is likely due to explicitly modeling physician-related variability.

**Conclusion:** Hierarchical models can yield better performance when there is physician-related variability as in the case of identifying relevant information in the EMR.

## 1 Introduction

A key source of frustration with electronic medical record (EMR) systems stems from the inability to retrieve relevant patient information efficiently [1, 2, 3, 4]. Current EMR systems do not possess sophisticated search capability nor do they prioritize patient information relative to the clinical task at hand [5, 6]. The inability to identify relevant patient information can lead to poor care and medical errors [7, 8, 9]. Further, in complex clinical environments, such as the intensive care unit (ICU), large quantities of data per patient accumulate rapidly [10], which can exacerbate information retrieval challenges. EMR systems that prioritize the display of relevant patient information are therefore needed to minimize the time and effort that physicians spend in identifying relevant information.

Various solutions have been proposed for effective prioritization and display of patient information in EMR systems [11, 12, 13, 14], most of which are based on rules that have been developed to customize and organize the display of patient information. In contrast to rule-based approaches, we developed and evaluated a data-driven approach called the learning EMR (LEMR) system in a prior study [15, 16]. The LEMR system tracks physician information-seeking behavior and uses it to learn machine learning models that predict which information is relevant in a given clinical context. Those predictions are used to highlight the relevant data in the EMR system to draw a physician’s attention.

However, information-seeking behavior has been shown to vary across individual physicians as well as across EMR system user types such as physicians, nurses, and pharmacists [1, 5]. In this study, we use hierarchical models to explicitly model this variability because such models have been shown to be useful when the data are collected from subjects with different behaviors [17]. In particular, we compare the performance of hierarchical logistic regression models and standard logistic regression models in predicting relevant patient information in a LEMR system.

The remainder of this paper is organized as follows. In the Background section, we review the LEMR system, briefly describe hierarchical models, and describe prior work on physician-related variability. In the Methods section, we describe the data collection and preparation, the experimental details, and the evaluation measures. We present the results of the experiments in the Results section, and close with Discussion and Conclusion sections.

## 2 Background

In this section, we provide brief descriptions of the LEMR system, hierarchical models, and past studies that have examined physician-related variability.

### 2.1 The LEMR system

The LEMR system uses a data-driven approach to prioritize patient information that is relevant in the context of a clinical task [15, 16]. The system uses machine learning to automatically identify and highlight relevant patient information for a specified task, for example, the task of summarizing a patient’s clinical status at morning rounds in the ICU. In ICU morning rounds, the clinical team reviews pertinent information and the status of each patient; for each patient, one team member reviews information in the EMR system and orally presents a summary of the patient’s clinical status to the team. Reviewing and identifying relevant patient information, called pre-rounding, is time-consuming and laborious. The goal of the LEMR system is to use machine learning to automatically identify and highlight the relevant information required for a given clinical task such as pre-rounding. The predictive models of the LEMR system are derived using the information-seeking behavior of physicians when they search for relevant information in the EMR in the context of the clinical task. In particular, eleven critical care physicians reviewed the EMRs of ICU patients and marked the information that was relevant to pre-rounding, and predictive models were developed from this data.

### 2.2 Hierarchical models

Hierarchical models, also known as *multilevel* models, are useful in modeling hierarchically structured data because they can capture variability at different levels of the hierarchy [17]. For example, consider predicting the mortality rate in a hospital with several units, such as critical care, general medical care, and emergency care. The data has a two-level hierarchical structure with the hospital at the first level and the units at the second level of the hierarchy. The overall mortality rate at the hospital level is obtained by combining the unit-level mortality rates in some fashion. A hierarchical model explicitly estimates the variability of the mortality rates across the units and uses those estimates to derive the hospital level mortality rate, which can result in a better estimate of the overall mortality rate compared to using non-hierarchical models.

In a similar fashion, the information-seeking data used to develop the LEMR models has a two-level hierarchical structure, where the top level corresponds to data that denote the entire *population* of physician reviewers and the bottom level corresponds to data that denote individual physicians. For specific patient information such as serum creatinine, its relevance is expected to differ across physician reviewers. A hierarchical model of the LEMR data explicitly captures this variability that is likely to be useful in deriving more accurate predictive models.

### 2.3 Physician-related variability

Physician-related variability in healthcare outcomes has been of interest for decades, going back to the 1970s with studies reporting the effects of geographic location on clinical outcomes such as mortality and length of stay [18]. In particular, variation in individual physician characteristics and practice styles has been recognized as a source of variability in clinical outcomes after adjusting for the health status of patients and the quality of healthcare services [5, 19, 20, 21, 22, 23, 24, 25]. For example, variability in cesarean section rates has been attributed to physician practice style after controlling for patient characteristics and risk factors, status of the medical facility, and physician years of experience [25]. A study concluded that variability across individual physicians may impact the quality of preference-sensitive critical care delivery [20]. A recent study analyzed physician search patterns in the EMR and uncovered considerable variation in information-seeking behavior [5]. In general, hierarchical modeling has been applied in various clinical settings to account for physician-related variability where the data has a hierarchical structure and can be grouped by a variety of factors such as country, state, or hospital site [26, 27, 28, 29, 30, 31, 32].

## 3 Method

In this section, we first describe the dataset and the data preparation steps. Then we describe the experimental methods including the development and evaluation of predictive models.

### 3.1 Dataset

One-hundred seventy-eight ICU patient cases with a diagnosis of either acute kidney failure (AKF; ICD-9 584.9 or 584.5; 93 cases) or acute respiratory failure (ARF; ICD-9 518.81; 85 cases) were selected randomly from patients who were admitted between June 2010 and May 2012 to an ICU at the University of Pittsburgh Medical Center. Eleven critical care medicine physicians reviewed the patient cases in the LEMR system and for each patient indicated which patient information was relevant to the task of pre-rounding in the ICU. The recruited reviewers included ICU fellows and attending clinicians from the Department of Critical Care Medicine at the University of Pittsburgh. Each physician reviewer was instructed to review up to 30 patient cases. The first four cases were the same for all reviewers and were used as burn-in cases; these cases were not included in the dataset. The remaining cases were different for each reviewer and each physician reviewed and annotated as many cases as they could during one to two sessions that lasted a total of four to six hours. Because the cases had some variation in the amount of patient information they contained and the physicians varied in the speed of reviewing, all physicians did not review the same number of cases.

The dataset consists of two sets of variables including the predictor variables (or predictors) and target variables (or targets) that we now describe in detail. Predictor variables include demographics, admitting diagnosis, vital signs, ventilator settings, input and output measurements, laboratory test results, and medication administration data. A few variables such as demographics and admitting diagnosis are static, that is, their values do not change during the ICU stay, while the remaining variables, which constitute the majority of the predictors, are temporal and have multiple values during the ICU stay. For example, age (in years) is a static predictor variable while blood urea nitrogen (BUN) is a temporal predictor variable as it is usually measured multiple times during an ICU stay.

Target variables include any data in the EMR, such as vital signs, ventilator settings, input and output measurements, laboratory test results, and medication administration data that a physician may annotate as relevant for the task of pre-rounding. A target variable can take either *relevant* or *not relevant* values. As an example, for a patient with AKF, BUN = *relevant* denotes that BUN was measured for the patient and was sought, found, and annotated by a physician as relevant. If BUN was measured for the patient but was not sought by a physician, then the target is denoted as BUN = *not relevant*. A target variable may be missing too; for example, when BUN is not measured for the patient, it would not be available for a physician to seek and find. We developed a predictive model for each target variable such as BUN that predicts whether it is relevant in a particular patient. To develop a BUN model, we used all predictor variables described in the previous paragraph and used only data in which the BUN target was not missing.

The difference between predictor and target variables is in the values they take; i.e., a target variable takes values of either *relevant* or *not relevant*, whereas a predictor variable’s values are the measured values that are recorded in the EMR. For example, when BUN is a predictor, it takes numeric values in milligrams per deciliter (mg/dL) unit^1^, whereas as a target variable, it takes a value of either relevant or not relevant. Consequently, a model for predicting whether or not BUN is relevant may contain numeric values for BUN as a predictor variable.

### 3.2 Data preparation

We transformed the dataset into a representation that is amenable to the application of machine learning methods. In particular, for each temporal predictor variable we generated between 4 to 36 features (feature expansion in Figure 1).

**Figure 1:**
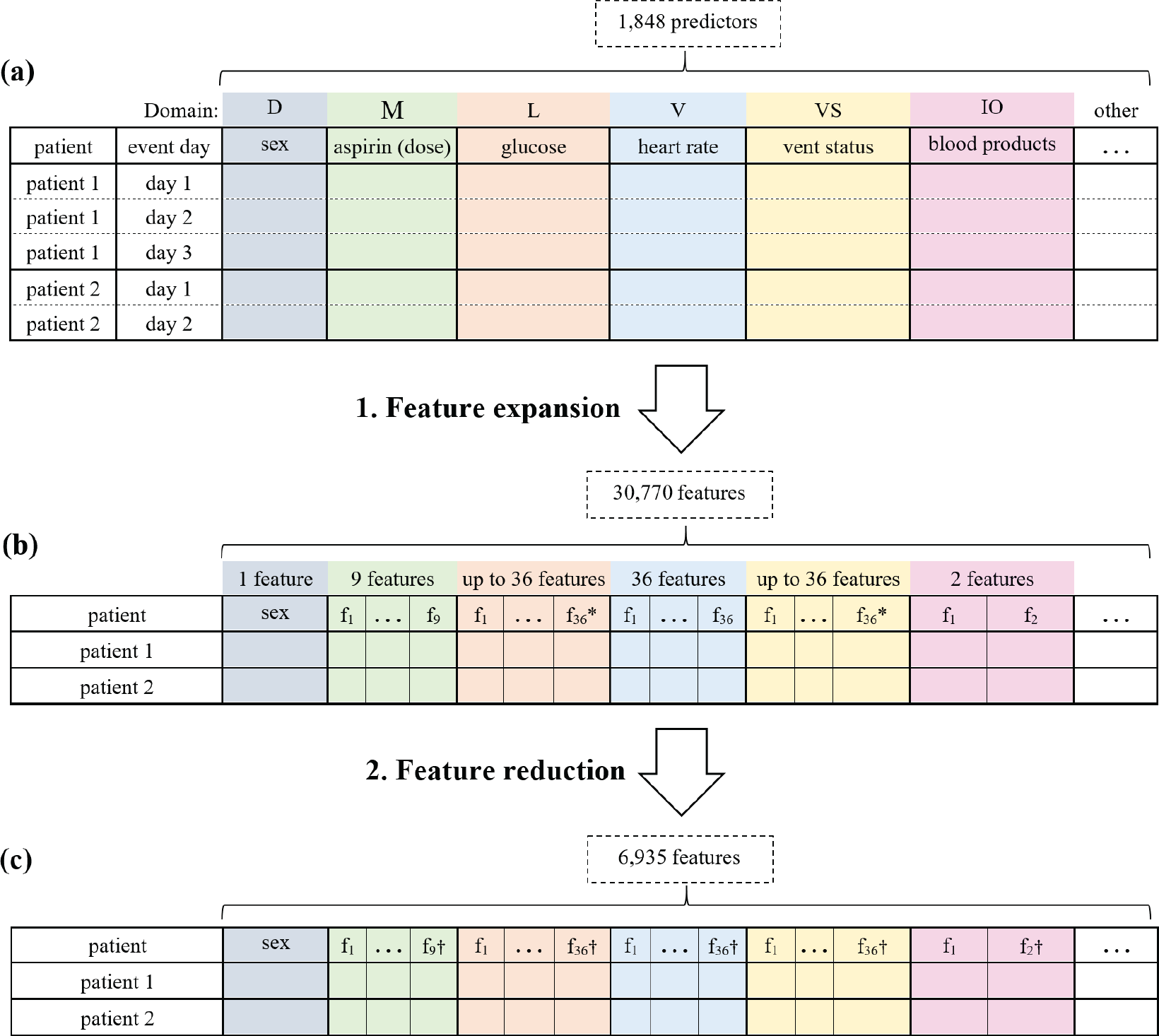
Steps in preparing the predictor variables. **(a)** presents the predictor variables for two example patients as measurements with one row per day. The colors represent data domains; D=demographics, M=medication administrations, L=laboratory test results, V=vital signs, VS=ventilator settings, IO=input/output, and other=other domains. **(b)** shows the result of expanding the temporal predictor variables (total = 1,848) to features (total = 30,770). This step flattens the data so that a patient that is represented by multiple rows is now represented by a single row. * denotes that the number of expanded predictors differs depending on the predictor value type (e.g., nominal or continuous). **(c)** shows the features after feature reduction, in which the number of features is reduced to 6,935s. † indicates that the number of features may be different for each variable in the domain.

The number of features for a temporal predictor was based on (1) the data domain of the predictor variable (e.g., medication administration or laboratory result) and (2) the type of the predictor variable (e.g., nominal or continuous). For example, for each medication variable we generated four features including an indicator of whether the drug is currently prescribed, the time elapsed between first administration and the current time, the time elapsed between the most recent administration and the current time, and the dose at the most recent administration. For each laboratory test result, vital sign, and ventilator setting, we generated up to 36 features including an indicator of whether the event or measurement ever occurred, the value of the most recent measurement, the highest value, the lowest value, the slope between the two most recent values, and 30 other features. More details on the feature expansion are given in [33].

The dataset consisted of 178 patient cases and 1,864 raw predictor variables. Feature expansion resulted in a total of 30,770 features. Since the dimensionality of the data was high, we reduced the number of features (feature reduction in Figure 1) by removing those features where the values were missing in every patient case, had the same value for every case (i.e., had zero variance), or the values were duplicates of another variable. Feature reduction resulted in a total of 6,935 features.

We selected as target variables 73 EMR data items that had been annotated as relevant (positive) in 9 or more patient cases. Table A1 in Appendix C contains the list of target variables along with the number of cases in which each target variable was relevant, as well as the number of cases where the target variable was available for selection (i.e., the value was not missing).

### 3.3 Experimental methods

#### 3.3.1 Predictive models

An HLR model is a generalization of a standard logistic regression model in which the data are clustered into groups and the model intercept and coefficients can vary by group [17]. Figure 2 shows the structure of a 2-level HLR model in which the LEMR data are clustered into groups of patient cases reviewed by each physician. Parameters at the lower level represent the physician-level models for the 11 physician reviewers, and parameters at the upper level represent the model for the entire population of physician reviewers (i.e., population-level model). For a more detailed description of HLR models see Appendix A.

**Figure 2:**
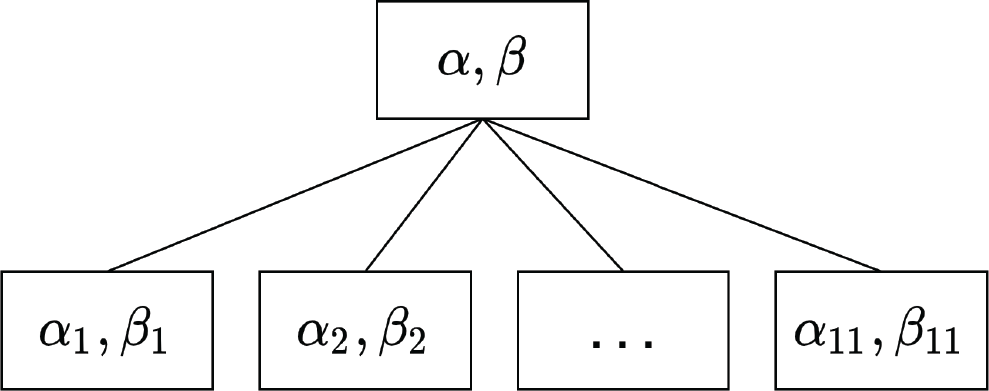
A 2-level HLR model for LEMR data. The lower level represents physician-level intercepts (*α*_*i*_) and coefficients (*β*_*i*_) where *i* = 1, …, 11 denotes the physician identifier. The upper level represents the intercept and coefficients (*α, β*) for the population-level model.

We developed HLR predictive models for each of the selected 73 targets. Each predictive model of a target variable is formulated as a binary classification problem where the model learns to identify cases in which the target variable is relevant. To investigate the utility of HLR over non-hierarchical models, we used LR as baseline models in which the physician identifier was included as an indicator variable. We implemented the HLR models using the brms package [34] in R, which uses No-U-Turn Sampler (NUTS) (as an extension of the Hamiltonian Monte Carlo algorithm) to estimate the posterior distribution of model parameters. In our experiments, we set the NUTS sampler to use 4 Markov chains; each chain included 400 iterations of sampling where the first 200 were used to calibrate the sampler. A total of 4 × 200 = 800 posterior samples for each HLR model parameter were obtained. LR models were implemented using the glmnet package in R [35].

#### 3.3.2 Cross validation

Each model was trained and evaluated independently in a stratified 10-fold cross validation setting. At each iteration of the cross validation, the patient cases were randomly split into a training set (9 folds) and a test set (1 fold), while preserving the original distribution of the target variable. Hyperparameter tuning and data preprocessing such as imputing missing values and feature selection were performed during cross validation. More details are described in Appendix B.

#### 3.3.3 Performance measures

We measured the predictive performance of each model with the area under the receiver operating characteristic (ROC) curve (AUROC), area under the precision-recall curve (AUPRC), and expected calibration error (ECE) [36]. AUROC is a measure of model discrimination and varies from 0.5 and 1, where 0.5 denotes an uninformative model and 1 represents perfect discrimination. AUPRC summarizes the precision-recall curve where precision (or positive predictive value) and recall (or sensitivity) values at different thresholds are plotted as a curve. The AUPRC varies from 0 to 1 is and is commonly used in binary classification problems when the data are imbalanced (i.e., when cases with one label are more prevalent than cases with the other label).

ECE is a measure of model calibration. In a perfectly calibrated model, outcomes with predicted probability correspond to a fraction of positive cases in the data. ECE is derived from the probability calibration curve [37] where the sorted predicted probabilities are partitioned into bins; in each bin *i*, calibration error is defined as the absolute difference between the mean of predicted probabilities (*p*_*i*_) and the fraction of positive outcomes (*o*_*i*_). ECE is the weighted average of the calibration errors over all bins:

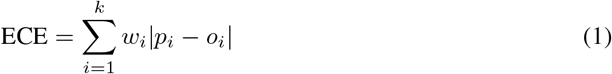

where *w*_*i*_ denotes the fraction of cases that fall into bin *i*. Lower ECE denotes a better calibrated model.

## 4 Results

We report the variability across the physician reviewers and then report the results of the predictive performance of LR and HLR models from three perspectives: *overall, per-target*, and *per-physician*. Table 1 summarizes the physician characteristics and the number of patients that each physician reviewed within the two diagnostic groups, AKF and ARF.

**Table 1:** Years of ICU experience for each physician and the number of patient cases each physician reviewed.

| Physician identifier | Years of ICU experience | # Cases reviewed (#ARF, #AKF) |
| --- | --- | --- |
| 1 | < 1 | 15 (8, 7) |
| 2 | 1 | 15 (10, 5) |
| 3 | 3 | 12 (5, 7) |
| 4 | < 1 | 17 (8, 9) |
| 5 | 1 | 15 (9, 6) |
| 6 | 1 | 15 (7, 8) |
| 7 | 2 | 22 (10, 12) |
| 8 | 1 | 20 (11, 9) |
| 9 | 1 | 16 (8, 8) |
| 10 | 2 | 16 (8, 8) |
| 11 | 7 | 15 (9, 6) |

### 4.1 Variability in information-seeking behavior

We define a descriptive statistic called *average relevance proportion* (ARP) to measure the information-seeking behavior of each physician reviewer. An ARP value for a physician is defined as the average proportion of EMR data items that the physician sought as relevant. We calculated the ARP values over the 73 EMR data items that were used as target variables. Figure 3 shows the physician ARP values separately for each of the diagnostic groups. Each circle denotes the ARP value for the corresponding physician on the x-axis and each error bar represents a 95% confidence interval (CI) for an ARP value. In the ARF diagnosis group, the ARP CIs for physicians 1, 7, and 8 do not overlap with those of the other physicians, which indicates a potential variability in information-seeking behavior between these physicians and the rest. Similar variability is observed in the AKF group, where the ARP CIs of physicians 1, 3, 7, and 8 differ from those of the other physicians.

**Figure 3:**
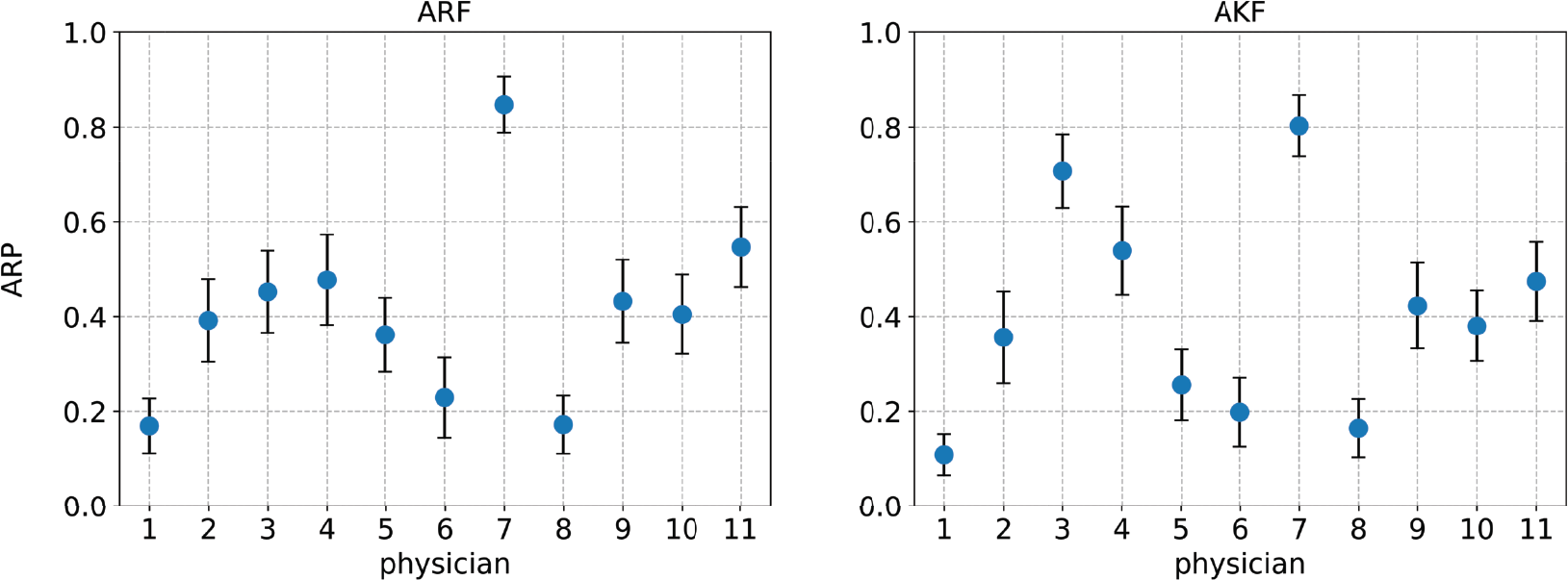
Per-physician ARP values over 73 target variables. A blue circle denotes the ARP value and an error bar denotes a 95% CI. The panel on the left is for ARF cases and the panel on the right is for AKF cases.

### 4.2 Overall performance

The overall performance of each model family (LR and HLR) was calculated by concatenating the predictions for all 73 target variables into a single vector and using that vector to compute the performance metrics. Table 2 reports the AUROC, AUPRC, and ECE for the LR and HLR models across all 73 target variables. For AUROC values, the 95% CI and p-value were calculated using Delong’s method [38, 39]. The 95% CI for AUPRC values was derived using the logit intervals method [40] and the p-value was calculated using the Wald z-test. For ECE values, we set *k* = 100 in Equation 1 and obtained a vector of 100 calibration errors to compute 95% CIs and a t-test p-value. Figure 4a shows the overall ROC and calibration curves for LR and HLR models. Note that for the calibration curves, we set the number of bins to *k* = 10 for better visibility.

**Table 2:** Overall AUROC, AUPRC, and ECE for LR and HLR models over all 73 target variables and across all physicians. Higher AUROC and AUPRC show better discrimination power while lower ECE denotes better probability calibration. The best values for each metric are in boldface.

| Measure | LR | HLR | p-value |
| --- | --- | --- | --- |
| AUROC | 0.75 (0.74-0.76) | **0.81 (0.80-0.82)** | < 0.001 |
| AUPRC | 0.665 (0.663-0.667) | **0.763 (0.762-0.765)** | < 0.001 |
| ECE | 0.16 (0.14-0.17) | **0.07 (0.06-0.08)** | < 0.001 |

**Figure 4:**
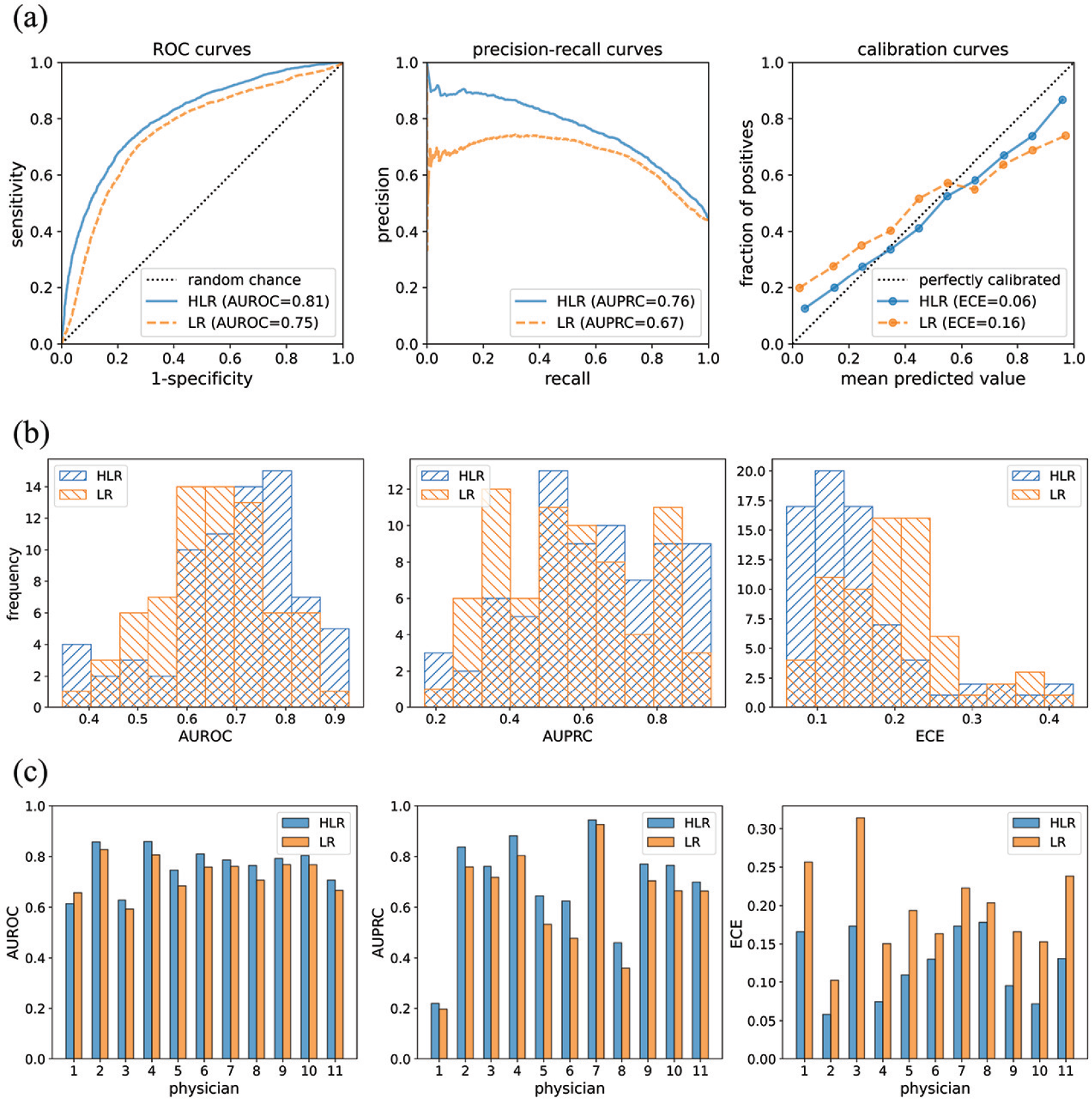
**(a)** ROC, precision-recall, and calibration curves over all 73 target variables across all physicians. For the calibration curves, the closer a curve is located to the dotted diagonal line, the more calibrated the corresponding approach is. **(b)** Distribution of AUROC, AUPRC, and ECE values for 73 models. Forward-slash hatches in blue represent the distributions for HLR models and backslash hatches in orange denote the distributions for LR models. The AUROC and AUPRC distributions for HLR models are right-skewed relative to the LR models, which show that HLR models generally have better discrimination power. The distribution of ECE values of HLR models is left-skewed relative to the LR models, which means that HLR models are generally better calibrated than LR models. **(c)** AUROC, AUPRC, and ECE values for each physician reviewer over all 73 models. The values for HLR models are shown in blue and the values for LR models are shown in orange. The AUROC and AUPRC values are higher for HLR models than for LR models, except for the AUROC value for physician 1. All the ECE values are lower for HLR models, which mean that HLR models are better calibrated than the LR models.

### 4.3 Per-target performance

For per-target performance, we computed the predictive performance for each target variable, which resulted in vectors of AUROC, AUPRC, and ECE values each with a length of 73, for each model family (LR and HLR). Distributions of per-target performance measures are shown as histograms in Figure 4b for each model family. Histograms of the two model families are overlaid for better comparison. Additional details are provided in Table A1 in Appendix C where AUROC, AUPRC, and ECE values are reported for each target variable.

### 4.4 Per-physician performance

For per-physician performance, we computed the predictive performance for each physician, which resulted in 11 AUROC, AUPRC, and ECE values for each model family (LR and HLR). Figure 4c presents the per-physician bar plots of the performance measure values; the bars for HLR and LR models are displayed side by side for better comparison. Per-physician calibration curves are presented in Figure A1 in Appendix C.

## 5 Discussion

Our results show that HLR models perform better than LR models when predicting which information a physician will seek in a future patient case. Moreover, the ECE results show that HLR models are generally better calibrated than LR models. In general, the more calibrated the probabilities are that are output by a predictive model, the higher the expected utility of the decisions that will be made using that model; in the case of the LEMR system, those decisions involve which information is worthwhile to highlight in the EMR of a given patient.

Although most physician reviewers had similar years of ICU experience, we observed a considerable degree of variability in information-seeking behavior across physicians in terms of ARP values. Because the study patients were selected to have a similar level of complexity, patient cases are unlikely to be the source of this variability. Controlling for physicians’ years of experience in LR models was not as effective in improving predictive performance as estimating individual physician variability using the HLR models. This shows the advantage of HLR models over standard models in the presence of unexplained variability.

The per-physician performance measures in Figure 4c show that HLR models learn physician-specific models that perform better in terms of both discrimination and calibration. Although HLR models fit a separate model for each physician, the inherent regularization in these models prevents overfitting. In particular, as population and physician-specific parameters are estimated at the same time, a pooling effect occurs that prevents a physician-specific model from overfitting when the sample size is small.

Furthermore, HLR models allow for a detailed investigation at the physician level because each physician model has its own set of parameters. Figure 5 demonstrates a few instances of the detailed information that can be obtained from an HLR model. Each panel in Figure 5 represents the distributions of a model parameter in an HLR model for each physician and for all physicians as a whole. Investigating the physician-specific parameters can lead to a better understanding of factors that influence a physician’s information-seeking behavior.

**Figure 5:**
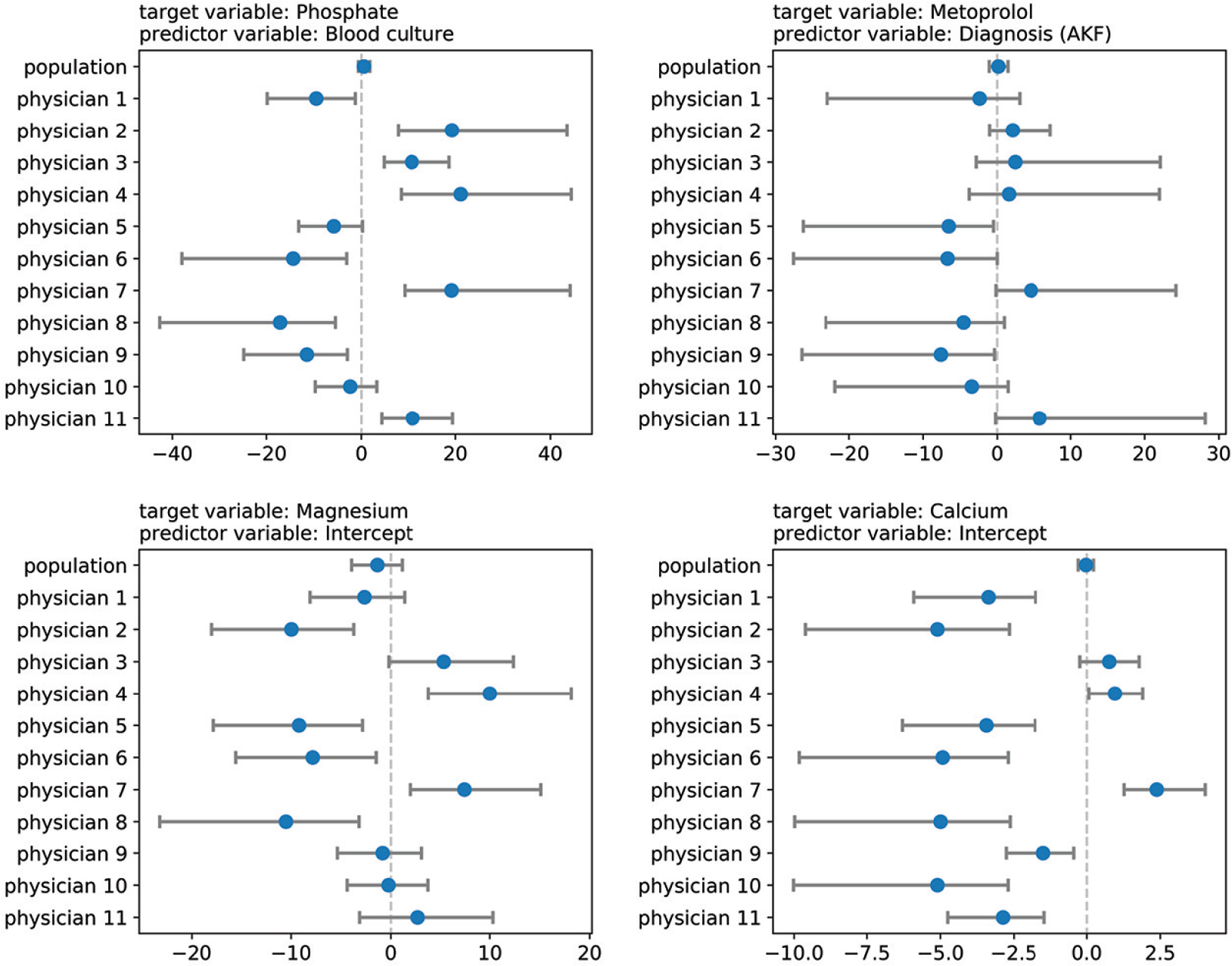
Examples of variation among physicians as seen from the values of the coefficients of a specific predictor variable. Each panel shows estimates of the coefficients of a predictor variable in an HLR model. A circle denotes the median value and the bar denotes the 80% credible interval for the posterior distribution of the model parameter.

## 6 Limitations

One limitation of this study was the relatively modest amount of annotated data. Having experts review and annotate data is an expensive and time-consuming task in many domains, especially in medicine. It takes many hours for a physician to review and annotate a small number of patient cases in the EMR, which makes it challenging to collect large amounts of annotated data in the LEMR system. Due to this limitation, the number of positive samples for most target variables was modest. As a result, we derived models for only 73 target variables out of 865 available target variables. Nevertheless, this restriction can be addressed by using scalable data collection methods. For example, a scalable solution based on eye-tracking technology has been proposed to automatically identify information that physicians seek in the EMR [41].

Another limitation of this study is that we did not model the reliability of the annotations across physicians. In practice, physicians will not agree on which information is relevant due to differences in knowledge, level of expertise, and subjective preferences. In a previous study, we observed that poor agreement among physicians on which information is relevant for the same case and clearly specified clinical task [15]. Moreover, inexperienced physicians may provide annotations that are judged to be erroneous by expert physicians. By pooling annotations from all physicians the HLR models are more influenced by physicians who have annotated a larger number of cases, and if disproportionally more cases are erroneously annotated in the training data, the models will provide poor recommendations. Thus, it is important to distinguish between correct and erroneous annotations which may be challenging in complex patient cases where even experts may disagree on which patient information is relevant. One approach to addressing this limitation is to develop a consensus model that physicians collectively converge to and combine such a model with physician-specific models [42].

Despite the advantage of HLR models in terms of performance, a drawback of HLR models is the added complexity due to the additional per-level parameters. This complexity creates new challenges in parameter estimation and interpretation. Compared to LR models, training HLR models requires more computing power and there are more hyperparameters to tune, including the choice of prior distributions.

## 7 Conclusion

Displaying large quantities of patient information in EMR systems with little prioritization can adversely influence the decision-making process of physicians and compromise the safety of patients. A data-driven solution was recently proposed as a learning EMR (LEMR) system that uses machine learning to identify and prioritize relevant data in the EMR for physicians. The current study improves the performance of LR models by using HLR models.

We trained 2-level HLR models that simultaneously learn physician-specific models at one level and a population model at another level. We evaluated the discrimination and calibration performance of HLR models in identifying relevant data items in the EMR. Our results show that HLR models perform significantly better than LR models. Moreover, we demonstrated that HLR models provide details about the physician-specific models that can be used to investigate physicians’ information-seeking behaviors in the EMR system.

## Data Availability

The dataset is not publicly available.

## 9 Acknowledgments

This research was supported in part by the University of Pittsburgh Center for Research Computing (CRC) through the resources provided. We specifically acknowledge the assistance of the research faculty consultants at CRC. The study was approved by the University of Pittsburgh IRB under protocol PRO14020588.

## Appendix

### A Hierarchical logistic regression (HLR) models

Hierarchical logistic regression (HLR) models are a generalization of standard logistic regression (LR) models in which distinct logistic regression models are fit at different levels of hierarchically structured data [17]. In the context of the LEMR system, we define an HLR model as follows (a boldface character represents a matrix or vector of parameters or values). Let *D* = {***X, y***} be a LEMR data set where ***X*** denotes a set of *N* patient cases in the EMR, each with *K* predictor variables including demographics, medication administrations, laboratory test results, and vital signs. Let ***y*** represent an EMR data item for *N* patient cases with binary values of *relevant* and *not relevant*. The data is reviewed by *J* physicians, each reviewing *n*_*j*_ cases such that 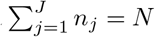. We formulate an HLR model as follows:

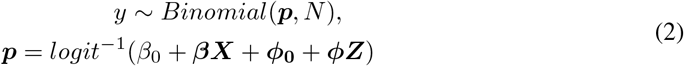

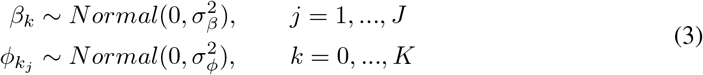

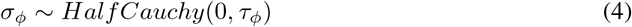

where *β*_0_ and ***β*** are population-level intercept and coefficients, and ***ϕ***_**0**_ and ***ϕ*** denote physician-level intercept and coefficients. ***Z*** corresponds to the physician-level design matrix, which is a sparse expansion of ***X*** with *N* rows and *J* × *K* columns, splitting ***X*** into *J* segments. Below is a simple example of ***X*** and ***Z*** matrices with *N* = 5 patients reviewed by *J* = 2 physicians (colored in blue and red) and *K* = 3 predictor variables:

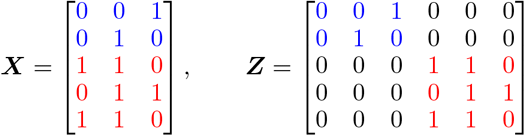

In Equation 3, *β*_*k*_ and *ϕ*_*k*_ denote the coefficients of the *k*^th^ predictor variable (*k* = 0 denotes the intercept) for the population and physician *j*’s models, and are assumed a priori to be centered at 0 with standard deviations *σ*_*β*_ and *σ*_*ϕ*_, respectively. We define the prior for *σ*_*ϕ*_ as a Cauchy distribution with scale parameter *τ*_*ϕ*_, and restrict it to positive values (hence the *half* Cauchy). *σ*_*β*_ and *τ*_*ϕ*_ are the model hyperparameters and are tuned in the model training phase. Markov chain Monte Carlo (MCMC) methods are used to estimate the parameters’ posterior distributions. MCMC methods draw samples sequentially from the posterior distribution and improve the draws at each step to better approximate the distribution.

### B Cross validation details

Each model was trained and evaluated independently in a stratified 10-fold cross validation setting. The following preprocessing steps were applied to the training and test sets at each iteration:

1. *Imputation*. Missing values in predictor variables were imputed by the median or mode for continuous and discrete variables, respectively. The imputed value was derived from the training set and applied to both the training and test sets.
2. *Feature selection*. We used supervised univariate feature selection to reduce the number of features before deriving models. In particular, we used analysis of variance (ANOVA) and Fisher’s exact tests (significance level = 0.01) for continuous and binary predictor variables, respectively. We limited the predictors to a maximum of 100 variables. The feature selection was performed on the training set and then applied to both the training and test sets.
3. *Feature standardization*. Continuous predictor variables were rescaled to be centered at zero and have a unit standard deviation. We calculated the mean and standard deviation statistics from the training set and used them to standardize both training and test sets.

Each model was trained on the training set and evaluated on the test set. Model hyperparameters were tuned in an inner stratified 3-fold cross validation of the training set. In particular, for HLR models, we selected the best values of the hyperparameters (i.e., *σ*_*β*_ and *τ*_*ϕ*_ in Equations 3 and 4 above) from the set of values {0.01, 0.1, 1, 5, 10}. For LR models, LASSO regularization was used and the optimal regularization parameter (*λ*) was chosen from an automatically generated sequence of 100 values as described in [35].

### C Tables and figures

**Table A1:** Performance of predictive models for 73 target variables. Higher AUROC and AUPRC denotes better discrimination and lower ECE denotes better calibration. Relevant/Available: number of cases in which the target variable was annotated as relevant over number of cases for which the target variable was measured in the EMR and was available for annotation. Domain: IO=input/output, L=laboratory test result, M=medication, V=vital sign, VS=ventilator setting.

| Target variable | Domain | Relevant/<br>Available | AUROC |  | AUPRC |  | ECE |  |
| --- | --- | --- | --- | --- | --- | --- | --- | --- |
|  |  |  | HLR | LR | HLR | LR | HLR | LR |
| acetaminophen | M | 12/68 | 0.67 | 0.51 | 0.31 | 0.22 | 0.14 | 0.2 |
| albumin | L | 19/107 | 0.86 | 0.84 | 0.49 | 0.54 | 0.07 | 0.08 |
| albuterol ipratropium | M | 15/59 | 0.71 | 0.87 | 0.49 | 0.77 | 0.15 | 0.08 |
| ALT (SGPT) | L | 23/104 | 0.78 | 0.71 | 0.57 | 0.45 | 0.11 | 0.17 |
| ammonia | L | 12/39 | 0.65 | 0.51 | 0.47 | 0.35 | 0.14 | 0.26 |
| ampicillin sulbactam | M | 9/20 | 0.57 | 0.48 | 0.55 | 0.6 | 0.39 | 0.47 |
| anion gap | L | 19/111 | 0.87 | 0.75 | 0.69 | 0.54 | 0.08 | 0.08 |
| aspirin | M | 15/45 | 0.63 | 0.7 | 0.54 | 0.71 | 0.13 | 0.17 |
| AST (SGOT) | L | 25/106 | 0.71 | 0.6 | 0.47 | 0.36 | 0.13 | 0.19 |
| band cell count | L | 13/81 | 0.69 | 0.72 | 0.38 | 0.39 | 0.13 | 0.09 |
| base solution | M | 50/81 | 0.63 | 0.62 | 0.73 | 0.65 | 0.2 | 0.22 |
| bicarbonate | L | 103/167 | 0.79 | 0.7 | 0.83 | 0.8 | 0.11 | 0.14 |
| bicarbonate (HCO3), arterial | L | 11/102 | 0.63 | 0.74 | 0.19 | 0.35 | 0.09 | 0.06 |
| bilirubin, direct | L | 16/82 | 0.75 | 0.6 | 0.64 | 0.27 | 0.1 | 0.16 |
| bilirubin, total | L | 36/103 | 0.82 | 0.78 | 0.65 | 0.66 | 0.13 | 0.17 |
| blood urea nitrogen (BUN) | L | 114/166 | 0.76 | 0.66 | 0.83 | 0.8 | 0.14 | 0.16 |
| calcium | L | 41/155 | 0.76 | 0.79 | 0.56 | 0.54 | 0.11 | 0.1 |
| central venous pressure (CVP) | V | 31/103 | 0.76 | 0.59 | 0.63 | 0.38 | 0.11 | 0.24 |
| chlorhexidine topical | M | 20/86 | 0.88 | 0.81 | 0.71 | 0.63 | 0.13 | 0.11 |
| chloride | L | 106/167 | 0.85 | 0.77 | 0.88 | 0.81 | 0.09 | 0.11 |
| dextrose 5% in water | M | 17/46 | 0.63 | 0.55 | 0.6 | 0.46 | 0.29 | 0.33 |
| docusate | M | 9/47 | 0.85 | 0.82 | 0.66 | 0.59 | 0.15 | 0.08 |
| famotidine | M | 26/77 | 0.75 | 0.7 | 0.57 | 0.55 | 0.12 | 0.19 |
| fentanyl | M | 18/83 | 0.66 | 0.59 | 0.41 | 0.39 | 0.14 | 0.22 |
| fraction of inspired oxygen (FiO2) | VS | 95/142 | 0.72 | 0.69 | 0.83 | 0.81 | 0.13 | 0.18 |
| furosemide | M | 28/66 | 0.65 | 0.66 | 0.57 | 0.56 | 0.15 | 0.25 |
| glucose | L | 114/164 | 0.66 | 0.64 | 0.8 | 0.81 | 0.17 | 0.2 |
| hemoglobin | L | 123/156 | 0.76 | 0.57 | 0.9 | 0.82 | 0.08 | 0.16 |
| heparin | M | 38/97 | 0.68 | 0.71 | 0.52 | 0.6 | 0.14 | 0.24 |
| hydrocortisone | M | 10/20 | 0.35 | 0.28 | 0.44 | 0.4 | 0.3 | 0.54 |
| input/output (I/O) | IO | 81/167 | 0.8 | 0.74 | 0.83 | 0.72 | 0.13 | 0.14 |
| INR | L | 62/118 | 0.7 | 0.65 | 0.7 | 0.63 | 0.1 | 0.22 |
| insulin aspartate (Novolog) | M | 11/27 | 0.36 | 0.36 | 0.34 | 0.36 | 0.45 | 0.45 |
| insulin glargine (Lantus) | M | 13/21 | 0.35 | 0.21 | 0.6 | 0.48 | 0.37 | 0.6 |
| insulin regular (Humulin R, Novolin R) | M | 36/77 | 0.61 | 0.55 | 0.65 | 0.56 | 0.21 | 0.34 |
| ionized Ca | L | 30/122 | 0.72 | 0.62 | 0.49 | 0.35 | 0.11 | 0.23 |
| lactate | L | 50/109 | 0.73 | 0.68 | 0.72 | 0.59 | 0.1 | 0.21 |
| lactulose | M | 9/16 | 0.92 | 0.67 | 0.95 | 0.74 | 0.19 | 0.33 |
| lansoprazole | M | 9/34 | 0.73 | 0.65 | 0.55 | 0.47 | 0.2 | 0.24 |
| levetiracetam | M | 9/16 | 0.93 | 0.79 | 0.94 | 0.88 | 0.24 | 0.22 |
| lorazepam | M | 9/37 | 0.35 | 0.43 | 0.34 | 0.26 | 0.34 | 0.25 |
| magnesium | L | 73/163 | 0.77 | 0.72 | 0.71 | 0.65 | 0.13 | 0.13 |
| metoprolol | M | 19/57 | 0.52 | 0.43 | 0.36 | 0.35 | 0.24 | 0.38 |
| metronidazole | M | 16/28 | 0.67 | 0.59 | 0.76 | 0.67 | 0.22 | 0.33 |
| midazolam | M | 9/51 | 0.62 | 0.59 | 0.3 | 0.3 | 0.14 | 0.19 |
| mixed venous oxygen saturation (SvO2) | V | 9/39 | 0.64 | 0.47 | 0.54 | 0.32 | 0.21 | 0.23 |
| mode | VS | 71/140 | 0.75 | 0.66 | 0.73 | 0.66 | 0.11 | 0.2 |
| neutrophils | L | 24/146 | 0.75 | 0.68 | 0.39 | 0.35 | 0.09 | 0.13 |
| norepinephrine | M | 17/35 | 0.51 | 0.47 | 0.48 | 0.56 | 0.25 | 0.35 |
| oxygen saturation (SaO2), arterial | V | 103/166 | 0.77 | 0.73 | 0.83 | 0.78 | 0.15 | 0.15 |
| pantoprazole | M | 16/42 | 0.79 | 0.81 | 0.73 | 0.69 | 0.15 | 0.15 |
| partial pressure of carbon dioxide (PaCO2), arterial | L | 31/130 | 0.69 | 0.64 | 0.41 | 0.32 | 0.09 | 0.21 |
| partial pressure of oxygen (PaO2), arterial | L | 30/129 | 0.85 | 0.7 | 0.68 | 0.41 | 0.08 | 0.16 |
| PEEP | VS | 9/22 | 0.84 | 0.87 | 0.8 | 0.87 | 0.26 | 0.17 |
| pH, arterial | L | 46/129 | 0.67 | 0.63 | 0.54 | 0.44 | 0.17 | 0.22 |
| phosphate | L | 69/160 | 0.77 | 0.74 | 0.75 | 0.62 | 0.13 | 0.19 |
| piperacillin tazobactam | M | 24/47 | 0.81 | 0.67 | 0.85 | 0.67 | 0.2 | 0.27 |
| platelets | L | 116/156 | 0.8 | 0.65 | 0.89 | 0.83 | 0.08 | 0.17 |
| potassium | L | 120/167 | 0.73 | 0.67 | 0.87 | 0.84 | 0.12 | 0.15 |
| potassium chloride | M | 28/125 | 0.84 | 0.78 | 0.66 | 0.58 | 0.07 | 0.09 |
| propofol | M | 17/42 | 0.45 | 0.5 | 0.49 | 0.45 | 0.34 | 0.35 |
| PTT | L | 15/101 | 0.57 | 0.63 | 0.24 | 0.25 | 0.13 | 0.16 |
| respiratory rate (RR) | V | 121/167 | 0.7 | 0.64 | 0.85 | 0.82 | 0.15 | 0.2 |
| senna | M | 10/43 | 0.8 | 0.85 | 0.55 | 0.55 | 0.17 | 0.12 |
| sodium (Na) | L | 128/167 | 0.72 | 0.57 | 0.89 | 0.81 | 0.11 | 0.24 |
| sodium chloride 0.9% | M | 65/144 | 0.62 | 0.57 | 0.56 | 0.53 | 0.19 | 0.28 |
| temperature | V | 144/167 | 0.72 | 0.57 | 0.91 | 0.88 | 0.09 | 0.18 |
| troponin | L | 10/60 | 0.48 | 0.42 | 0.17 | 0.16 | 0.23 | 0.22 |
| tube status | VS | 38/123 | 0.62 | 0.59 | 0.52 | 0.4 | 0.17 | 0.15 |
| vancomycin | M | 36/71 | 0.61 | 0.54 | 0.64 | 0.53 | 0.22 | 0.31 |
| vancomycin, trough | L | 13/41 | 0.46 | 0.62 | 0.37 | 0.48 | 0.28 | 0.19 |
| ventilator status | VS | 15/124 | 0.87 | 0.83 | 0.63 | 0.52 | 0.07 | 0.05 |
| white blood cell count (WBC) | L | 132/156 | 0.72 | 0.59 | 0.91 | 0.87 | 0.09 | 0.14 |
| Average |  |  | 0.7 | 0.64 | 0.62 | 0.56 | 0.16 | 0.21 |

**Figure A1:**
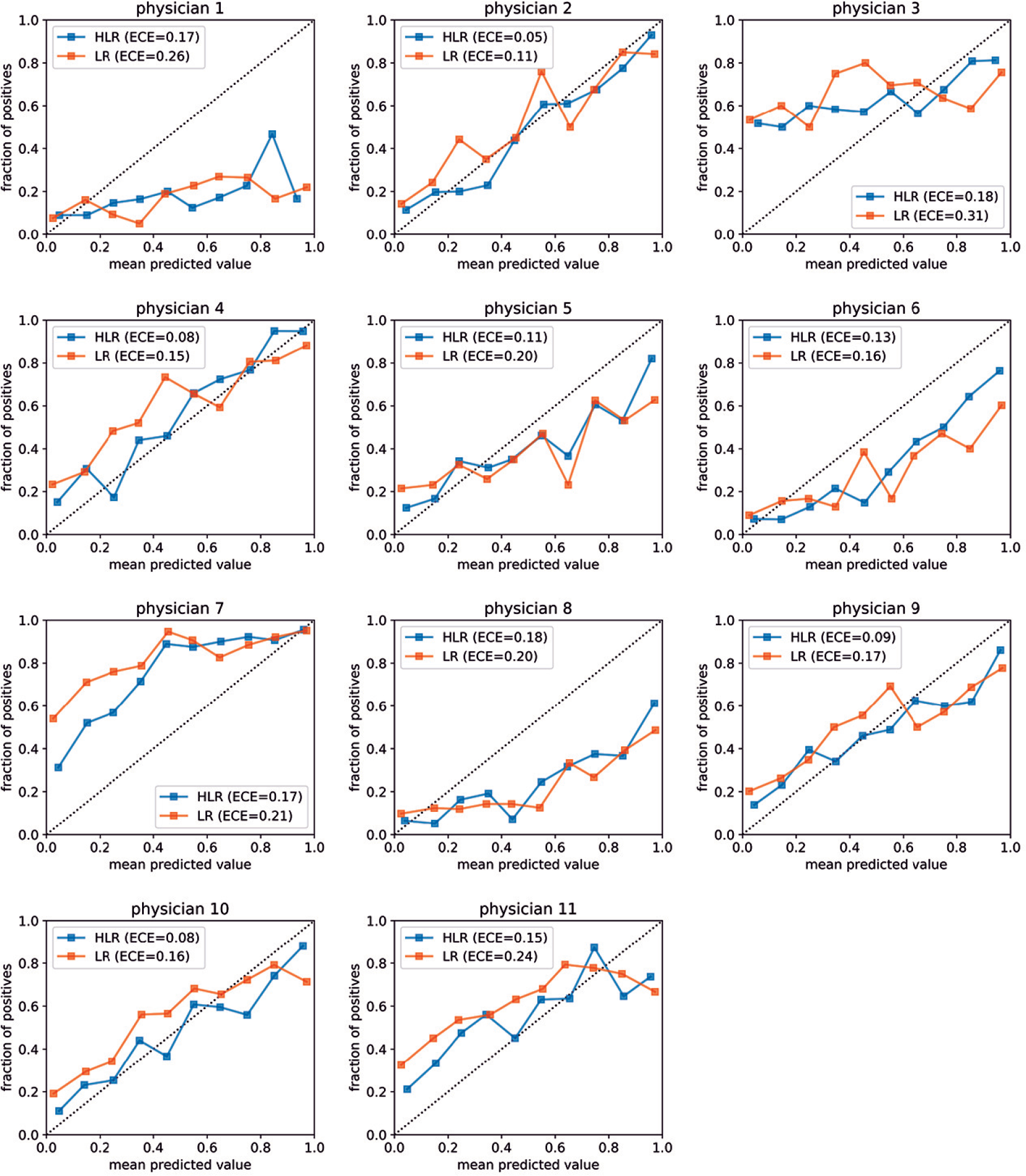
Calibration curves for per-physician models. Based on ECE values, HLR models are better calibrated than LR models for all physicians.

## Footnotes

1 Note that BUN may have been measured multiple times for a patient case and therefore, take several numeric values. We summarize these values as a fixed-length vector as described in the Data preparation section.

